# Antibody response after a single dose of BBV152 vaccine negatively correlates with pre-existing antibodies and induces a significant but low levels of neutralizing antibodies to Omicron variant

**DOI:** 10.1101/2022.02.07.22270612

**Authors:** Suman Das, Janmejay Singh, Heena Shaman, Balwant Singh, Anbalagan Anantharaj, Patil Sharanabasava, Rajesh Pandey, Rakesh Lodha, Anil Kumar Pandey, Guruprasad R. Medigeshi

## Abstract

Most adults in India have received at least one dose of COVID-19 vaccine and also been infected naturally during the pandemic. As immunization of individuals continues under this situation where the virus has attained endemicity, we assessed whether this hybrid immunity is further boosted by a single dose of BBV152, an inactivated SARS-CoV-2 vaccine, and, if these antibodies can neutralize SARS-CoV-2 delta and omicron variants. We found that natural infection during the second wave in 2021 led to generation of neutralizing antibodies against other lineages of SARS-CoV-2 including the omicron variant, albeit at a significantly lower level for the latter. A single dose of BBV152 boosted antibody titers against the delta and the omicron variants but the antibody levels remained low for the omicron variant. Boosting of antibodies showed negative correlation with baseline neutralizing antibody titers suggesting anergy of the immune system in individuals with high levels of antibodies.

## INTRODUCTION

Serosurveillance studies have showed that 69% of the Indian population had antibodies for COVID-19 after the second wave^1^ and a subsequent serosurvey from Delhi showed seropositivity of over 90%^2^ suggesting that India was/is a heterogenous mix of people with immunity to COVID-19 due to vaccination or natural infection or both after the second wave. As COVID-19 has attained a state of endemicity and majority of the population got vaccinated after the second wave, there have been no studies to measure the impact of pre-existing humoral immunity on subsequent vaccination. As antibody levels wane and the risk of reinfection with the same variant and or a new variant warrants booster vaccination, it is imperative to understand the efficiency of boosting vis-à-vis pre-existing antibody levels. We enrolled subjects who got vaccinated with BBV152 as part of routine vaccination program and assessed the effect of pre-existing antibody levels on boosting after receiving a single dose of vaccine. We found that majority of the individuals had neutralizing antibodies to delta variant and natural infection with delta variant led to generation of neutralizing antibodies against other lineages of SARS-CoV-2 including the omicron variant although the level of antibodies was significantly lower for this new variant of concern as compared to delta and ancestral virus.

## METHODS

### Human Ethics

The study was approved by the Institutional ethics committees for human research at ESIC Hospital and Medical College and THSTI. Informed consent was obtained from all the participants.

### Human Samples

#### BBV152 cohort

An informed consent was obtained in person from each participant before they were being recruited in this study at the ESIC Medical College & Hospital, Faridabad between May – August 2021. All adults of > 18 years of age who came for the first dose of vaccine were eligible to participate in this study including those who had recovered from the COVID-19 in the recent past. In addition, participants were requested to provide any history related to COVID-19 infection. Blood samples of the participants were collected in anti-coagulant free vacutainers with the help of a professional phlebotomist and stored at 2 - 8 ºC. First blood sample (5 ml) was collected on day 1 prior to the administration of the first dose of the vaccine. Second blood sample (5 ml) was collected during the follow-up visit before the administration of the second dose of the vaccine.

### Cells

Vero E6 cells were obtained from European Collection of Authenticated Cell Cultures and maintained in Minimal essential medium (MEM) (Gibco) supplemented with 10% heat-inactivated fetal bovine serum (FBS), 100U of penicillin and 100 μg of streptomycin and L-glutamine (PSG) (Gibco), 1X non-essential amino acid mix (NEAA) (Gibco), 25 mM HEPES in 5% CO2 incubator. Calu-3 cells (Human lung epithelial cells derived from adenocarcinoma: ATCC-HTB-55) were maintained in Dulbecco’s minimal essential medium (DMEM) (HiMedia) supplemented with 10% heat-inactivated fetal bovine serum (FBS), 100U of penicillin and 100 μg of streptomycin and L-glutamine (PSG) (Gibco), 1X non-essential amino acid (NEAA) (Gibco),

### Viruses

SARS-CoV-2 to B.6 and delta lineage virus isolation has been described earlier^3–5^. SARS-CoV-2 Omicron isolate (sub-lineage BA.1) was obtained from Leo Poon^6^. SARS-CoV-2 variants were propagated in Vero E6 cells or Calu-3 cells^7^ and virus passaging was limited to four passages. All virus stocks used in this study was verified by whole genome sequencing using total RNA sample of the culture on Nanopore sequencing platform as described previously to confirm the variant^8^.

### Quantitative Nucleoprotein ELISA

The bacterial expression plasmid pET-28a(+) containing the codon optimized nucleocapsid (N) gene from severe acute respiratory syndrome-related coronavirus 2 (SARS-CoV-2), Wuhan-Hu-1 (GenBank: MN908947) having N-terminal hexa-histidine affinity purification tag was requested from BEI resources (NR-53507). His-tagged N protein was purified by Ni-NTA chromatography as per previous report ^9^. 96-well MaxiSorp ELISA plates (Nunc) were coated with 1 μg/mL purified N protein diluted in 1X PBS pH 7.4 and the plates were incubated for 1 h at room temperature (RT). Serum samples were inactivated using Triton X-100 (Sigma) to a final concentration of 1 % at RT for 1 h. The serum samples were two-fold serially diluted starting from 1:100 to 1: 6400 and 100 μl/well was added to the antigen-coated plate. After 30 min at room temperature, the plate was washed using 1X PBST (phosphate-buffered saline with 0.1 % Tween 20). After washing, 50 μl/well HRP-conjugated anti-human IgG was added and incubated at room temperature for 30 min. 100 μl/well of TMB substrate was added for 10 min and the reaction was terminated using 1M H_2_SO_4_ as stop solution. The intensity of the color was quantified by measuring absorbance in a microplate reader at 450 nm with 630 nm as reference wavelength. Antibody concentrations were calculated for each sample dilution by interpolation of the OD values on the 4-parameter logistic (4-PL) standard curve from in-house reference control (calibrated as a secondary standard using WHO reference standard reagent (20/130)) and adjusted according to their corresponding dilution factor using Gen5 software. The assay has a limit of quantitation of 3 binding antibody units/mL (BAU/mL). This assay has been validated in-house and accredited under ISO17025:2017 standard.

### Quantitative RBD ELISA

Recombinant spike protein receptor binding domain (RBD) ELISA was performed as described earlier^4,10^. Recombinant spike protein Receptor Binding Domain (RBD) antigen of SARS-CoV-2 were coated onto 96-well polystyrene plate (0.1μg/well) and incubate coated plate at 4°C for 18-22 hours. Antigen-coated plates were washed with wash buffer and incubated by adding 200 μl of blocking buffer (5% non-fat dry milk powder in PBST). Serum samples were inactivated by adding 10 μl of 10% Triton X-100 in 90 μl serum to obtain a final concentration of 1% Triton X-100. Samples were gently mixed and incubated at room temperature for 1 hour. After washing the plate with wash buffer, 100 μl of diluted serum (1:50-1:6400 diluted in blocking buffer) was added to each well and incubated at RT (23±2°C) for 30 ± 10 min. Substrate was added and OD was recorded and data was analyzed as described in the previous section. The assay has a limit of quantitation of 12 binding antibody units/mL (BAU/mL). This assay has been validated in-house and accredited under ISO17025:2017 standard.

### Virus microneutralization assay

Virus microneutralization assay by focus reduction neutralization titer assay using indicating virus isolates was performed as described earlier with minor modifications in omicron variant staining^7^. Briefly, serum samples were serially diluted from 1:20 to 1:640 and virus neutralization was tested in Vero E6 cells. Cells were incubated for 24 hours for ancestral (B.6) and Delta (B.1.617.2) variants and for 32 hours for Omicron (B.1.1.529) variant. After incubation, cells were fixed with formaldehyde solution and then stained with anti-spike RBD rabbit polyclonal antibody at 1:2000 dilution (Sino Biologicals, Cat. No. 40592-T62) for 1 h, followed by HRP-conjugated anti-rabbit antibody at 1:4000 dilution (Invitrogen, Cat. No. G-21234) for 1 h. For Omicron isolate, incubation was extended to 32 h and a 1:1000 dilution of anti-nucleocapsid primary antibody (Genscript, Cat. No. A02048-1) and 1:500 dilution of HRP-conjugated goat anti-mouse IgG secondary antibody (Invitrogen, Cat. No. A16072) was used for staining. Cells were washed with PBS and incubated with TrueBlue substrate (KPL inc, USA, Cat. No. 5510-0030) for 10 minutes and washed with sterile MilliQ water. Microplaques developed after staining were quantified by AID iSPOT reader (AID GmbH, Strassberg, Germany). The raw data generated from the AID iSpot Analyser in a 96-well format is pasted in a pre-defined protocol template for calculation of FRNT_50_ by using log_10_ transformed dilution value and neutralization percentages in an XY format. The Point-to-Point curve fit using a linear equation to fit each pair of data points was used to calculate the FRNT50 value. 50% neutralization values were calculated using SoftMax Pro GxP software v7.7.1 (Molecular Devices).

### Statistical analysis

Data was analysed and final graphs were prepared using GraphPad Prism (Version 9) software. Statistical significance was estimated by two-tailed, non-parametric Mann-Whitney test or Wilcoxon signed rank test as indicated.

## RESULTS

We collected baseline blood sample at the time of vaccination and a follow-up sample four weeks later, before the second dose in the months of June-July 2021. A total of 94 (37 females) subjects were enrolled. Median age of the subjects was 31.5 yrs (range: 18-67 yrs). Sixty-seven of 94 (71.3%) subjects were positive in a quantitative RBD-ELISA in the baseline sample. Four samples were indeterminate. We also measured the levels of nucleocapsid (N) antibodies by quantitative ELISA. Fifty five of the 94 (58.5%) samples were positive for N antibodies suggesting exposure in the second wave. After one dose of vaccination, positivity in RBD-ELISA increased to Eighty four of 94 samples (89.4%) and for N-ELISA 79 out of 94 samples (84%) were positive. The GMT of RBD antibodies increased significantly from 109 (95% CI: 76, 156) to 206 (95% CI: 163, 260) in baseline seropositive (Figure 1A) subjects, however, the increase in the GMT of N antibodies was from 18 (95% CI: 12, 25) to 25 (95% CI: 20, 31) which was not significant (Figure 1B).

**Figure 1:**
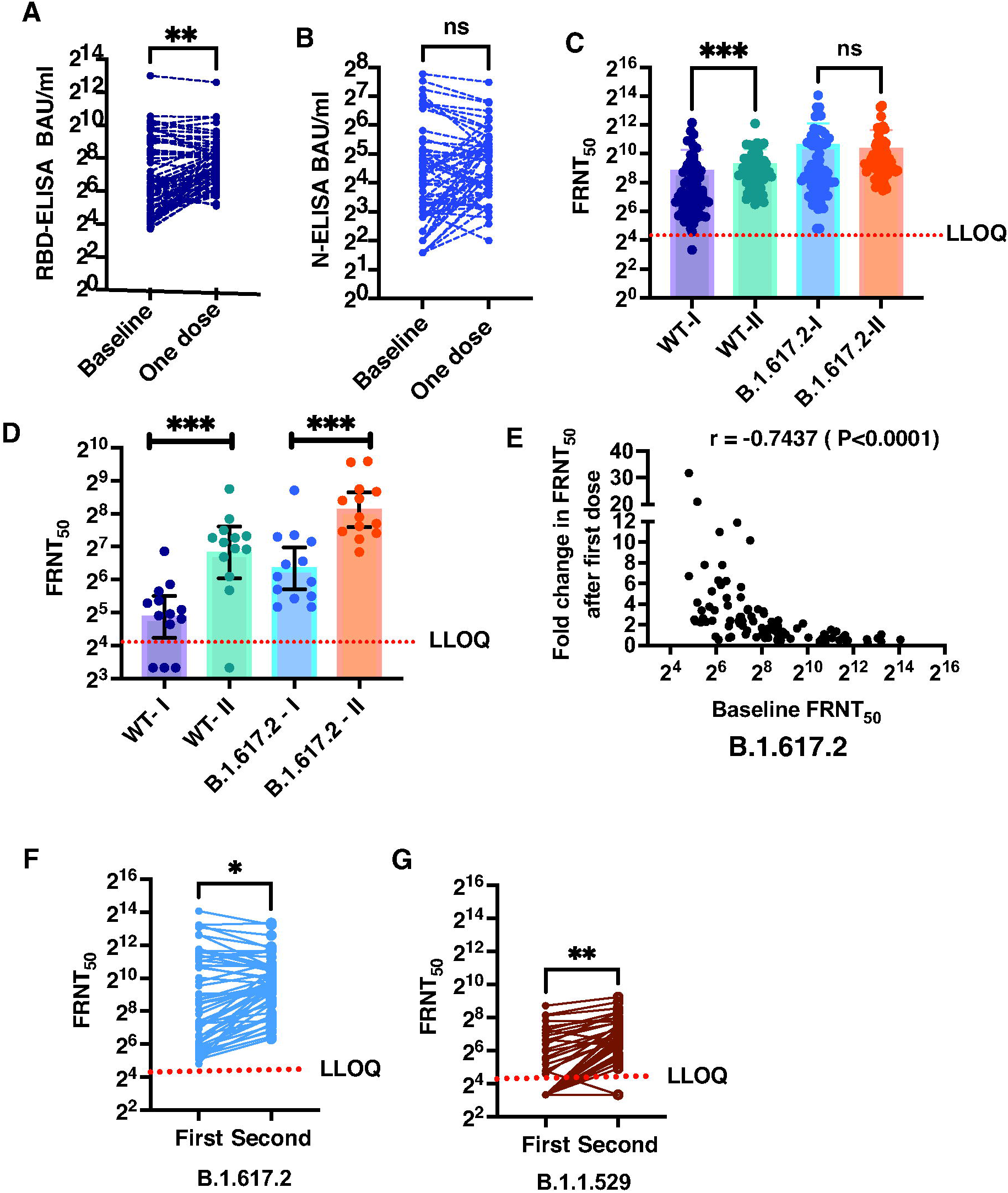
Antibody boosting after a single dose of BBV152 vaccination. Serum levels of antibodies against (A) SARS-CoV-2 RBD and (B) N protein was estimated by quantitative ELISA after a single dose of BBV152 vaccine in baseline and post-vaccination samples collected 3-4 weeks later. Data was expressed as Binding Antibody Units (BAU)/ml (international units). (C) FRNT_50_ titers in RBD ELISA-positive samples for the ancestral B.6 lineage (WT) and the delta variant (B1.617.2) in baseline and post-single dose vaccination samples. (D) FRNT_50_ titers in RBD ELISA-negative samples for the ancestral B.6 lineage (WT) and the delta variant (B1.617.2) in baseline and post-single dose vaccination samples. (E) Spearman correlation (r) between baseline FRNT_50_ titers and fold-change observed after single-dose of BBV152. (F and G) FRNT_50_ titers for the delta variant (B1.617.2) and omicron (1.1.529) variant in baseline and post-single dose vaccination samples. LLOQ: Lower limit of quantitation of the assay. Two-tailed t test P values are indicated by - * p<0.05; ** p<0.01; *** p < 0.001; **** p < 0.0001; ns - not significant.

We next measured the virus neutralizing antibody titers against the SARS-CoV-2 B.6 lineage virus from 2020^4^ (designated as WT) and B.1.617.2 (Delta variant) isolates in both the baseline and post-single dose vaccination samples by focus reduction neutralization titer (FRNT) assay. Overall GMT of neutralizing antibodies increased after one dose of BBV152 vaccination for both WT virus and delta variant (Table 1). Both RBD and N antibody levels correlated with neutralization titers against the delta variant after the first dose (Supplementary Figure S1). The FRNT data were further analyzed based on the seropositivity status at baseline in RBD-ELISA and we found that one dose of BBV152 vaccine led to significant increase in GMT for neutralizing antibodies against the WT virus (Table 1 and Figure 1C). However, the GMT for delta variant showed a marginal and insignificant increase from 531.9 (95% CI: 371.6, 761.2) to 783.2 (95% CI: 624.8, 981.7) in the RBD-ELISA positive subgroup at baseline (Table 1 and Figure 1C). Twenty-two out of 23 baseline RBD-ELISA negative samples showed the presence of neutralizing antibodies for WT and delta variant in FRNT assay, however, the levels of these antibodies were about 7-8 fold lower than that in the ELISA-positive samples (Table 1) suggesting that the antibody levels were below the level of detection of ELISA and these are not seronegative subjects. In samples collected from baseline RBD-ELISA negative participants, the GMT of antibodies against both the WT and B.1.617.2 lineage viruses increased significantly after receiving the first dose of the vaccine (Table 1 and Figure 1D). Overall, we observed that samples with FRNT_50_ >1000 failed to show any induction in antibodies after the first dose. We observed a clear negative correlation between the baseline FRNT_50_ titer for delta variant and the fold change in titer values after a single dose of vaccination (Figure 1E).

**Table 1:** Virus neutralization titers post single-dose of BBV152 vaccination based on RBD-ELISA positivity.

|  | Geometric Mean FRNT50 (95% CI) |  |  |  |  |  |
| --- | --- | --- | --- | --- | --- | --- |
| Sampling time | Baseline<br>**WT**<br>(A) | One dose<br>**WT**<br>(B) | P value<br>comparing<br>the titers<br>within each<br>group;<br>before and<br>after 1 dose<br>of vaccine<br>(A vs B)<br>(**Wilcoxon**<br>**signed**<br>**rank test**) | Baseline<br>**B.1.617.2**<br>(C) | One dose<br>**B.1.617.2**<br>(D) | P value<br>comparing<br>the titers<br>within each<br>group;<br>before and<br>after 1<br>dose of<br>vaccine (C<br>vs D)<br>(**Wilcoxon**<br>**signed**<br>**rank test**) |
| All (n=94) | 108.6<br>(81.3 –<br>145.1) | 256.4<br>(196.1 –<br>335.3) | <0.0001 | 291.4<br>(209.6 – 405.0) | 508.1<br>(398.9 –647.2) | 0.0126 |
| Groups based on<br>RBD ELISA<br>results |  |  |  |  |  |  |
| 1. RBD-positive at<br>baseline<br>(n=67) | 182.8<br>(133.6 –<br>250.0) | 422.2<br>(340.5 –<br>523.5) | 0.0001 | 531.9<br>(371.6 – 761.2) | 783.2<br>(624.8 –<br>981.7) | 0.448 |
| 2. RBD-Negative<br>at<br>baseline<br>(n=23) | 26.50<br>(19.8 – 35.5) | 57.32<br>(33.0 – 99.6) | 0.0033 | 63.94<br>(46.4 – 88.1) | 146.6<br>(95.7 – 224.7) | 0.0002 |
| P value<br>comparing the<br>titers between 2<br>groups based on<br>RBD ELISA<br>(using **Wilcoxon**<br>**rank sum test**) | <0.0001 | <0.0001 |  | <0.0001 | <0.0001 |  |

Comparison of titers at baseline and 3-4 weeks after 1<sup>st</sup> dose was performed using Wilcoxon signed rank test. Comparison of titers between the groups (based on RBD ELISA results) was done using Wilcoxon rank sum test.

While this study was in progress, omicron (B.1.1.529) emerged as a variant of concern and antibodies from most vaccines showed reduced efficiency in neutralizing this variant^7,11,12^. We randomly selected 55 paired samples (which had a FRNT_50_ value for the delta variant) to test for their ability to neutralize the omicron variant. Only twenty out of 55 baseline samples had detectable levels of neutralizing antibodies against omicron. By assigning a FRNT_50_ value of 10 for the samples which had no detectable levels of antibodies in the starting dilution (1:20) of the assay, we obtained a GMT of 22 (95% CI: 16, 31) for these 55 samples. This value was 18-fold lower than the GMT of delta variant which was 404 (95% CI: 248, 658). After a single dose of BBV152, the number of samples positive for neutralizing antibodies against omicron increased to thirty six out of 55 subjects with a significant increase in GMT (p = 0.0011) to 52 (95% CI: 36, 75), however, this was still 15-fold lower than the GMT for delta variant which was 784 (95% CI: 575, 1068) (Figure 1F and 1G). Whether the modest but significant boosting observed for omicron by BBV152 vaccination in seropositive individuals is protective or not warrants further studies from the ongoing booster vaccination campaign across the country.

Vaccination in early convalescent individuals led to poorer boosting as observed in other studies^13–15^. In naturally infected individuals, boosting after six months is predicted to increase the vaccine effectiveness against variants of concern^16^. We were not able to ascertain the exact date of past infection in most of these individuals which is not unusual as most COVID-19 infections are asymptomatic or mild and are not diagnosed. Nucleocapsid antibodies are known to decay with a half-life of 68 days^17^ and 59% of our study participants were positive for N antibodies suggesting that they could have been infected with SARS-CoV-2 within the past 6-8 months. Therefore, we cannot rule out the possibility that some of the subjects with high antibody titers were in their early convalescence and therefore, the boosting effect may not have been significant due to anergy of the immune system as has been observed in other studies^18^. Antibody boosting correlated negatively with the levels of pre-existing antibodies which indicates that a strategy is needed to prioritize high-risk individuals based on their antibody levels for booster vaccination. Nevertheless, the level of neutralizing antibodies for omicron variant after boosting in both seropositive and seronegative individuals remained much lower compared to the ancestral virus or the delta variant which is consistent with recent reports for CoronaVac, an inactivated vaccine^19^. However, the participants of our study were all non-vaccinated individuals and we cannot rule out the possibility that vaccination of these individuals after further reduction in antibody titers would have induced a better antibody response. It has been estimated that neutralizing antibodies are a good correlate of protection and contribute to about 60% of the protective efficacy of a vaccine which indicates that the cellular responses play a critical synergistic role along with antibodies in mediating protection from SARS-CoV-2 VoCs^4,5,16^. Some of the recent studies have also shown that heterologous boosting mounts a robust immune response to VoCs^20^. As many new vaccines are likely to be licensed in the coming months in India, more studies are required to measure the efficacy of homologous vs heterologous boosting against new variants of concern.

### Data availability

All the data are presented in this manuscript. Detailed Methods are available as online methods.

## Data Availability

All data produced in the present work are contained in the manuscript.

## ACKNOWLEDGEMENTS

We thank all the members of the bioassay lab for technical support. We thank Neha Garg and Shamsher Singh for data management. We thank all the participants who consented to enrol into the study.

## AUTHOR CONTRIBUTIONS

JS, HS, BS, AA, and PS performed experiments and analyzed the data. SD and AKP coordinated the study at clinical site, generated all the clinical site data and contributed reagents. RP sequenced the virus isolates and analyzed the data. RL provided critical inputs in experimental design, data analysis and writing the manuscript. GRM and AKP conceived the study, designed the experiments and analyzed the data. GRM wrote the manuscript. All authors have reviewed and approved the final version of the manuscript.

