## Supplementary Figure S1 for "Antibody response after a single dose of BBV152 vaccine negatively correlates with pre-existing antibodies and induces a significant but low levels of neutralizing antibodies to Omicron variant"

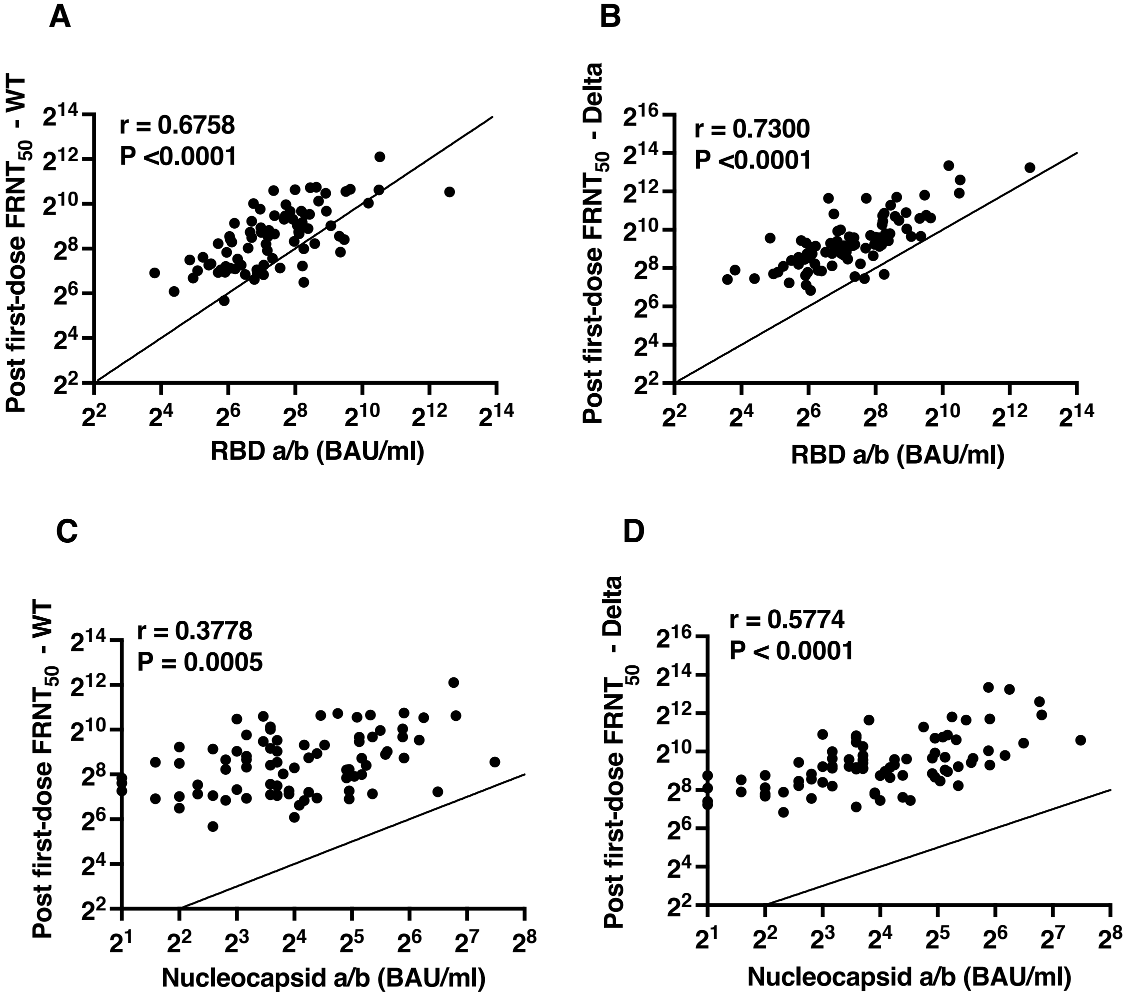


**Supplementary Figure S1: Neutralization antibody titers by FRNT correlate with RBD and N antibody titers by ELISA.** Spearman correlation (r) between RBD-ELISA binding antibody units (BAU/ml) and post-vaccination FRNT50 titers against the (A) B.6 lineage (WT) or the (B) Delta variant virus. Spearman correlation (r) between N-ELISA binding antibody units (BAU/ml) and post-vaccination FRNT50 titers against the (C) B.6 lineage (WT) or the (D) Delta variant virus. Two-tailed t test P values are indicated.
